# Symptomatology associated with the diffusion of the SARS-CoV-2 Lambda variant in Peru: An infodemiologic analysis

**DOI:** 10.1101/2021.08.24.21262245

**Authors:** Brandon Michael Henry, Maria Helena Santos de Oliveira, Thaís Barbosa de Oliveira, Kin Israel Notarte, Giuseppe Lippi

## Abstract

The SARS-CoV-2 (Severe Acute Respiratory Syndrome Coronavirus 2) Lambda variant rapidly diffused across Peru following its identification in December 2020, and had now spread worldwide. In this study, we investigated infodemiologic trends in symptomatology associated with the Coronavirus Disease 2019 (COVID-19) following the spread of SARS-CoV-2 Lambda variant in Peru, enabling infodemiologic surveillance of SARS-CoV-2 in regions with high circulation of this new variant. Weekly Google Trends scores were obtained for key symptom keywords between March 1^st^, 2020 and July 4^th^, 2021, whilst case count data were obtained from Peruvian Ministry of Health. Multiple time series linear regression was used to assess trends in each score series, using the week of December 27^th^ as cutoff for emergence of the Lambda variant. The significance of such trends was tested for each time period, before and after the cutoff date. A total 2,075,484 confirmed SARS-CoV-2 infections in Peru in relation to Google Trends data were analyzed. After Lambda variant emergence, searches for “diarrhea” demonstrated a change from a negative to positive correlation with weekly case counts and anticipated dynamic changes in case counts by 1-5 weeks. Searches for “shortness of breath” and “headache” remained consistently positively correlated to weekly case counts before and after Lambda emergence. No changes in searches for other common cold symptoms were observed, while no specific trends were observed for “taste loss” or “smell loss”. Diarrhea, headache, and shortness of breath appear to be the most important symptoms for infodemiologic tracking the current outbreak in Peru and other regions with high circulation of SARS-CoV-2 Lambda variant.

## Introduction

The rapid emergence of novel variants of the Severe Acute Respiratory Syndrome Coronavirus 2 (SARS-CoV-2) with potentially greater infectivity, transmissibility, severity, reduced neutralizing potential and/or ability to escape immune response poses a significant threat to global public health, especially in the face of continuing vaccine shortages.^1^ In December 2020, a new SARS-CoV-2 variant (lineage C.37) has been originally identified in Lima, Peru which has then been classified as “Lambda” by the World Health Organization (WHO).^2^ Early data suggest that this variant may have greater infectivity and capacity for immune escape from neutralizing antibodies from both naïve infection and vaccination, while changes in clinical severity remain to be determined.^3–5^ The lambda variant was classified in June 2021 by the WHO as “variant of interest”.^6^

Over the course of the ongoing Coronavirus Disease 2019 (COVID-19) pandemic, we have shown that infodemiology, which utilizes the volume of Google web searches for specific COVID-19 symptoms (*i*.*e*. keywords), is effective and reliable for predicting regional epidemiological trends^7,8^ and anticipating demand for SARS-CoV-2 testing^9^, assuming that symptomatology (*i*.*e*. the set of symptoms characteristic of a medical condition) of any emerging variants in the region remain consistent over time. In Peru, the Lambda variant accounted for 0.5% of cases in December 2020, then rapidly increased to 20.5% of cases in January 2021, 36.4% in February 2021, 79.2% in March 2021, up to 96.6% in April 2021.^2^ This dramatic trend in Peru presents the opportunity to investigate symptom keyword trends associated with case counts before and after emergence of this variant, given the homogenous spread within the country as opposed to other geographical regions which have experience the emergence and introduction of multiple variants over time. Here, we investigated infodemiologic trends in symptomatology associated with COVID-19 with the spread of the Lambda SASR-CoV-2 variant in Peru, in order to enable infodemiologic virus surveillance in regions with high circulation of this variant.

## Methods

Weekly Google Trends scores (Google Inc., Menlo Park, California, United States) were obtained for each symptom keyword between March 1^st^, 2020 and July 4^th^, 2021, for the geographic location Peru, which has approximately 31 million inhabitants and Spanish as its official language. The keywords searched were ‘fever’, ‘cough’, ‘shortness of breath’, ‘headache’, ‘smell loss’, ‘taste loss’, ‘fatigue’, ‘diarrhea’, ‘vomiting’, ‘nausea’, ‘nasal congestion’, ‘muscle pain’ and ‘stuffy nose’ in Spanish. A single unit in Google Trends reflects the relative search interest per week based on a 100-point scale, where the maximum value is established by the highest search volume for a particular term in the period studied. Weekly confirmed SARS-CoV-2 case counts in Peru were obtained from the Peruvian Health Ministry’s Open Data Repository.^10^ The case counts were then tabulated together with the Google Trend scores for the study period. Segmented linear regression for time series was used to assess trends in each score series, using the week of December 27^th^ as cutoff for emergence of the SARS-CoV-2 Lambda variant, and the significance of these trends was tested for each time period, before and after the cutoff date. Trend coefficients, confidence intervals and *p*-values are presented, as well as visual representation of fitted models. A *p*-value <0.05 was considered significant. To verify the stationarity (mean, variance and autocorrelation structure that do not change over time) before applying the regression in the scores, the Phillips-Perron (PP) and Kwiatkowski-Phillips-Schmidt-Shin (KPSS) tests were performed. To the first test, *p*-value <0.05 indicate significant stationarity and, to the second, *p*-value >0.05. To verify the presence of autocorrelation after applying the regression, the Durbin-Watson test was used. When the autocorrelation was significant, the adequacy of the model was used using the Cochrane-Orcutt method. To understand the relationship between case counts and Google Trend scores, time lagged cross correlations were calculated.

This study was performed in conformation with the Declaration of Helsinki, under the terms of applicable local legislation. The analysis was conducted solely based on the searches of unrestricted, publicly available databases; thus, no informed consent or institutional review board approval were required.

## Results

This study analyzed 2,075,484 total confirmed cases of SARS-CoV-2 infection in Peru in relation to Google Trends data throughout the study period (March 1^st^, 2020 to July 4, 2021). During this period, the number of new weekly SARS-CoV-2 cases ranged from 8 in the week of March 1^st^, 2020, to 63,558 in the week of April 4^th^, 2021. A total of 1,013,930 cases were observed in the months before December 27^th^, 2020, and 1,061,554 cases after that following emergence of the Lambda variant. Two major peaks in cases were observed in Peru, a pre-Lambda peak the first in the week of August 16^th^, 2020 and a Lambda peak in the second week of April 4^th^ 2021.

Google trends data and case counts for each week over the study period for a given symptom keyword are presented in Figure 1. The weekly score series for “vomiting” and “diarrhea” both showed significant positive trends between March 1^st^, 2020 and December 27, 2020 (*p*<0.001; *p*<0.01, respectively), followed by significant negative trends (*p*<0.001; *p*<0.05, respectively) after that time period, indicating that search interest in those topics had been rising since the early weeks of the pandemic, reaching a peak around the end of 2020 and declining afterward. The scores for “diarrhea” had significant and negative cross-correlation coefficients for weekly cases before the cutoff date, at no time lag as well as 1 through 5 weeks lag, meaning that higher scores were more likely to be followed by lower case counts. After the cutoff, the lagged “diarrhea” score series had mostly positive cross-correlations with the weekly case series, thus indicating a change in dynamics, where higher scores were more likely to be followed by higher case counts. The scores for “vomiting” also had mostly negative cross-correlations with the weekly case series before December 27^th^, but no significant cross-correlations after that date, thus signifying that higher scores were likely to be followed by lower case counts before the cutoff date, but had no significant association with case counts afterward.

**Figure 1.**
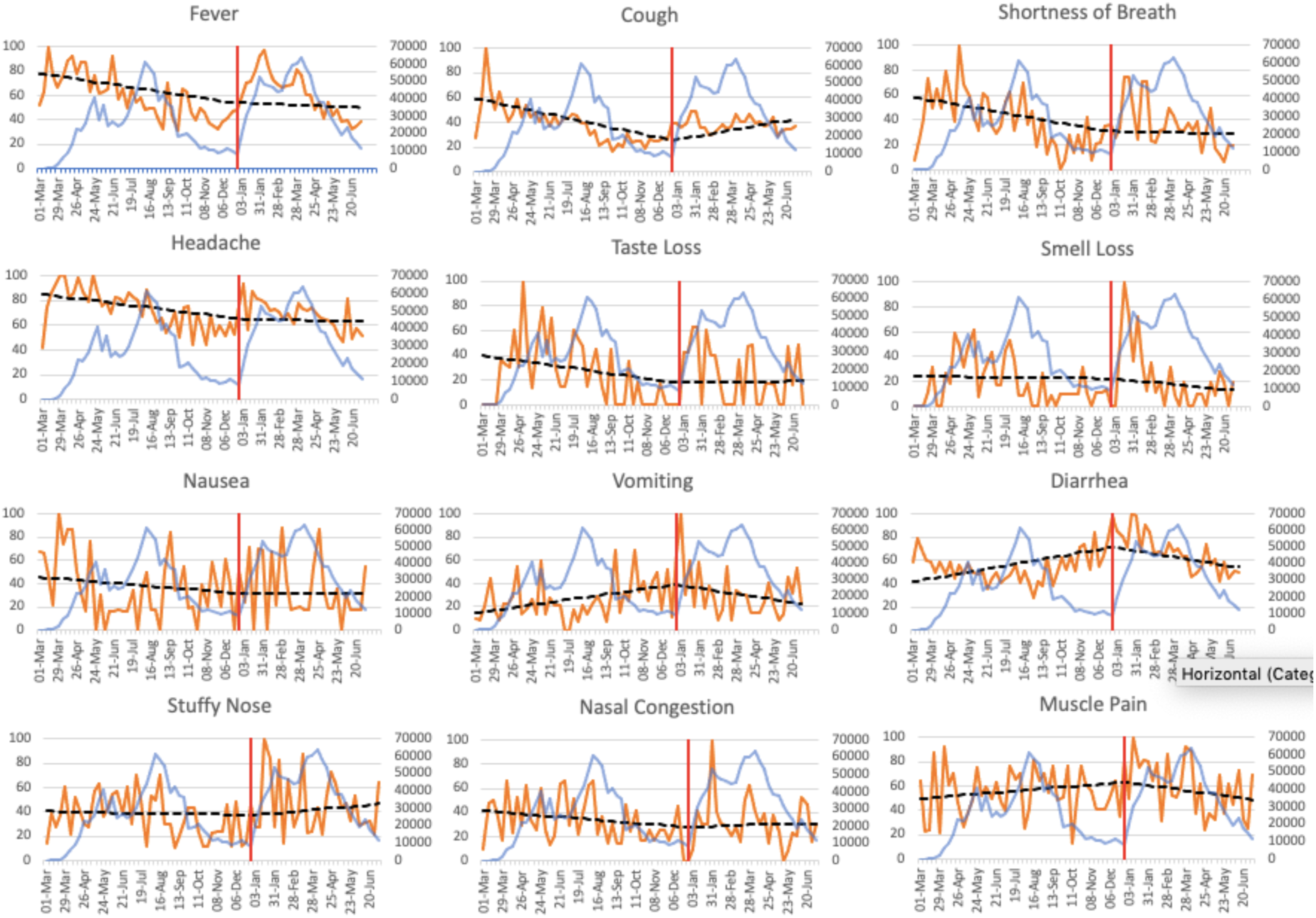
Weekly SARS-CoV-2 cases in Peru and Google Trend Scores for specific symptom keywords for searches conducted within the country from March 1^st^, 2020, to July 4^th^, 2021. Blue line represents number of weekly confirmed SARS-CoV-2 cases reflected by the left-sided y-axis; Orange line represents weekly Google Trend score reflected by the right-sided y-axis; Dashed black line represents overall search trend before and after December 27^th^, 2020 which is demarcated by a red vertical line. Not shown is the plot for fatigue which had no significant trends with weekly case counts.

The weekly score series for “cough” showed significant negative trend (*p*<0.001) before the end of 2020, and a significant positive trend (*p*<0.001) after that, indicating that Google search interest in the keyword had been declining since the beginning of the pandemic, but has been showing a significant increase since the cutoff date of December 27^th^. However, the scores had no significant cross-correlations with the weekly case series, thus indicating that no significant association of higher or lower scores with the number of cases in following weeks could be seen. Both “headache” and “shortness of breath” also had their peak Google search scores at the beginning of the pandemic, showing significant decline during the year 2020 (*p*<0.01; *p*<0.05, respectively), but no significant trend after December 27^th^. Both terms also had mostly positive cross-correlations with the weekly case series, both before and after the cutoff date, thus indicating that higher scores for these terms have been consistently associated with higher case counts in the following weeks.

The pathognomonic COVID-19 symptoms of “smell loss” and “taste loss” showed relative stability, with no significant trend variations before or after December 27^th^. Finally, no significant trends for fever were observed at any time point in the study period, while other general symptoms of upper respiratory tract infections, such as “fatigue” (PP test *p*<0.01, KPPS test *p*>0.05), “nasal congestion” (PP test *p*<0.01, KPPS test *p*>0.05), “stuffy nose” (PP test *p*<0.01, KPPS test *p*>0.05), and “muscle pain” (PP test *p*<0.01, KPPS test *p*>0.05), all showed stationary behavior before applying linear regression. After applying the regression, the results did not show statistically significant trends of change, which was expected, considering the mean, variance and covariance of any stationary series do not change over time. Full results of the time series analysis are presented in Table 1, and cross-correlation coefficients in Table 2.

**Table 1.** Time Series Analysis results for key COVID-19 symptoms before and after emergence of the lambda SARS-CoV-2 strain in Peru.

| Search Term | PP test | KPSS test | Linear tendency before December 27 | p-value (Linear regression) | Trend | Linear tendency after December 27 | p-value (Linear Regression) | Trend |
| --- | --- | --- | --- | --- | --- | --- | --- | --- |
| Fever | >0.05 | <0.05 | -0.57 (-1.38; 0.23) | >0.05 |  | 0.44 (-1.33; 2.22) | >0.05 |  |
| Fatigue | **<0.01** | **>0.05** | 0.26 (-0.12; 0.65) | >0.05 |  | -0.73 (-1.70; 0.22) | >0.05 |  |
| Headache | >0.05 | <0.05 | -0.45 (-0.71; -0.19) | **<0.01** | ↓ | 0.37 (-0.26; 1.02) | >0.05 |  |
| Nausea | >0.05 | <0.05 | -0.32 (-0.86; 0.21) | >0.05 |  | 0.30 (-1.01; 1.62) | >0.05 |  |
| Vomiting | >0.05 | <0.05 | 0.57 (0.20; 0.93) | **<0.001** | ↑ | -1.18 (-2.08; -0.29) | **<0.001** | ↓ |
| Diarrhea | >0.05 | <0.05 | 0.70 (0.19; 1.21) | **<0.01** | ↑ | -1.36 (-2.53; -0.18) | **<0.05** | ↓ |
| Smell Loss | >0.05 | <0.05 | -0.03 (-0.80; 0.73) | >0.05 |  | -0.31 (-2.09; 1.46) | >0.05 |  |
| Taste Loss | >0.05 | <0.05 | -0.52 (-1.20; 0.15) | >0.05 |  | 0.60 (-1.0; 2.20) | >0.05 |  |
| Cough | >0.05 | <0.05 | -0.78 (-1.11; -0.45) | **<0.001** | ↓ | 1.43 (0.67; 2.20) | **<0.001** | ↑ |
| Shortness of Breath | >0.05 | <0.05 | -0.65 (-1.23; -0.06) | **<0.05** | ↓ | 0.58 (-0.80; 1.96) | >0.05 |  |
| Stuffy Nose | **<0.01** | **>0.05** | -0.09 (-0.50; 0.33) | >0.05 |  | 0.44 (-0.58; 1.47) | >0.05 |  |
| Nose Congestion | **<0.01** | **>0.05** | -0.31 (-0.69; 0.07) | >0.05 |  | 0.45 (-0.48; 1.38) | >0.05 |  |
| Muscle Pain | **<0.01** | **>0.05** | 0.32 (-0.09; 0.73) | >0.05 |  | -0.85 (-1.86; 0.16) | >0.05 |  |

\* PP - Phillips-Perron test: a *p*-value below 0.05 indicates stationary behavior; KPSS - Kwiatkowski-Phillips-Schmidt-Shin test: a *p*-value above 0.05 indicates stationary behavior.

**Table 2.** Cross-correlation coefficients for Google Trends scores series and weekly case counts, before and after December 27^th^, 2020.

| Time lag (weeks) | Headache |  | Cough |  | Diarrhea |  | Shortness of Breath |  | Vomit |  |
| --- | --- | --- | --- | --- | --- | --- | --- | --- | --- | --- |
|  | Before | After | Before | After | Before | After | Before | After | Before | After |
| **0** | 0.102 | **0.364** | -0.14 | 0.281 | **-0.565** | 0.226 | 0.214 | **0.384** | -0.202 | -0.277 |
| **-1** | 0.16 | 0.357 | -0.044 | 0.224 | **-0.551** | 0.312 | **0.333** | **0.391** | **-0.334** | -0.168 |
| **-2** | 0.24 | 0.351 | 0.032 | 0.127 | **-0.493** | 0.364 | **0.352** | **0.378** | -0.248 | -0.008 |
| **-3** | **0.335** | 0.255 | 0.145 | -0.032 | **-0.484** | **0.386** | **0.371** | 0.279 | **-0.306** | -0.079 |
| **-4** | **0.387** | **0.397** | 0.157 | -0.166 | **-0.362** | **0.482** | **0.395** | 0.213 | **-0.321** | 0.256 |
| **-5** | **0.387** | 0.313 | 0.241 | -0.313 | **-0.339** | **0.449** | **0.358** | 0.185 | -0.268 | 0.246 |

\* Values represent linear regression coefficient and 95% CI.
PP - Phillips-Perron test: a *p*-value below 0.05 indicates stationary behavior; KPSS - Kwiatkowski-Phillips-Schmidt-Shin test: a *p*-value above 0.05 indicates stationary behavior.

## Discussion

Multiple previous studies, either by our group or by other teams which have investigated the trends in online search patterns within the context of regional COVID-19 outbreaks, have consistently shown that the frequency of Google searches of pathognomonic and non-pathognomonic (*i*.*e*., general) symptom keywords and SARS-CoV-2 surveillance data have a strong relationship, through which their temporal association enables predicting future trajectories of epidemic based on internet search volumes in a period of time, as well as the demand for SARS-CoV-2 testing and test positivity.^7–9^ However, this depends on consistency of symptoms over time and may also be influenced by emergence of novel SARS-CoV-2 variants with distinctive biological and clinical characteristics (*i*.*e*., higher virulence and/or pathogenicity, immune evasion, greater stability, and so forth).

In this investigation, we explored the symptomatology associated with diffusion of the SARS-CoV-2 Lambda variant in Peru, using a validated infodemiologic approach. A few interesting observations were found. First, shortness of breath is the most consistent symptom associated with all the major variants circulating in the region during the study period as demonstrated by positive search correlations with case counts before and after Lambda variant emergence. Second, a significant change in behavior was observed for search data on diarrhea after emergence of the Lambda variant. Prior to December 27^th^ 2020, negative correlations with case counts were observed but, after this date, searches for diarrhea positively correlated and anticipated spikes and drops in weekly case counts by 1-5 weeks. This suggests that diarrhea may be a more prominent feature, and perhaps early indicator of major circulation of the Lambda variant. No significant association was found with other gastrointestinal symptoms (*i*.*e*., nausea, vomiting) after Lambda variant emergence. Third, other generalized symptoms of upper respiratory tract infection, such as fever, cough, nasal congestion, fatigue, and muscle pain remained relatively stable over the study period, showing no change in their overall search trends following introduction of Lambda variant. Thus, we found no data to support that this emerging variant is more frequently presenting with common cold type of illness. Importantly, headache showed a rather consistent tracing across the study period, with positive search correlations, similar to shortness of breath, to weekly case counts. This is in line with previous findings in regard to the importance of headache in COVID-19 symptomatology.^11^. Finally, the pathognomonic COVID-19 symptoms of “smell loss” and “taste loss” displayed relative stability and no significant trend to suggest a positive correlation of these symptoms with the Lambda variant. This is in contrast to previous results published by our team, in which we have shown strong association between SARS-CoV-2 incidence and anosmia or dysgeusia^7,8^

Since being first identified in Peru in late 2020, this new SARS-CoV-2 lambda variant has now spread to countries across South America, with cases now being detected all around the world.^12^ Among the many mutations associated with the lambda variant, a novel spike protein mutation (L452Q) within the receptor binding domain may offer greater capability to bind to the virus human host receptor (Angiotensin Converting Enzyme 2 (ACE2)) and escape neutralizing antibodies produced from naïve infection, vaccination, or therapeutic antibody cocktails.^3^ Another seven-amino-acid deletion in the N-terminal domain of spike protein (*i*.*e*., RSYLTPGD246-253N) seems to confer a substantial resistance to vaccine-induced neutralization, which may favor the major spread of Lambda variant among COVID-19 vaccine recipients, especially in those with lower vaccine-triggered immunogenicity.^13^ Therefore, further studies would be urgently needed to investigate the real-world clinical and public health implications of the lambda variant.

This study is limited by several factors. Though Spanish is the most spoken language in Peru, it is just one of many potentially limiting the search. Furthermore, limitations to internet access, especially in rural regions of the country may partially bias the results. Seasonal cold and influenza patterns with similar baseline symptoms could also in part bias the results. Finally, increasing familiarity with key COVID-19 symptoms among the general public, especially pathognomonic symptoms like loss of taste and smell, could be confounding our analysis. Nonetheless, this would have important implications for the use of such symptoms to infodemiologically monitor the evolving pandemic.

## Conclusions

Following emergence of the SARS-CoV-2 Lambda variant in Peru, searches for diarrhea significantly increased and anticipated the dynamic changes in weekly case counts. Diarrhea, along with headache and shortness of breath, appear to be the most important infodemiologic symptoms for tracking the current outbreak in Peru and other regions with high Lambda variant circulation. On the other hand, common cold symptoms appear to be less frequently associated with searches in a region with high circulation of Lambda variant, while similarly no trend was observed for loss of taste or smell. These results provide some insight into symptomatology of SARS-CoV-2 Lambda variant, and highlight the key search terms that should be used for infodemiologic surveillance of SARS-CoV-2 in regions with high circulation of this variant.

## Data Availability

All data are reflected in the manuscript.

## Notes

### Competing Interest Statement

The authors have declared no competing interest.

### Funding Statement

No external funding was received.

### Author Declarations

The study was performed in conformation with the Declaration of Helsinki, under the terms of applicable local legislation. The analysis was conducted solely based on the searches of unrestricted, publicly available databases; thus, no informed consent or institutional review board approval were required.

